# A Deep Recurrent Reinforced Learning model to compare the efficacy of targeted local vs. national measures on the spread of COVID-19 in the UK

**DOI:** 10.1101/2021.05.21.20248630

**Authors:** Tim Dong, Umberto Benedetto, Shubhra Sinha, Arnaldo Dimagli, Massimo Caputo, Gianni D Angelini

## Abstract

**Objectives:** We have developed a deep learning model that provides predictions of the COVID-19 related number of cases and mortality in the upcoming 5 weeks and simulates the effect of policy changes targeting COVID-19 spread.

**Methods:** We developed a Deep Recurrent Reinforced Learning (DRRL) based model. The data used to train the DRRL model was based on various available datasets that have the potential to influence the trend in the number of COVID-19 cases and mortality. Analyses were performed based on the simulation of policy changes targeting COVID-19 spread, and the geographical representation of these effects.

**Results:** Model predictions of the number of cases and mortality of COVID-19 in the upcoming 5 weeks closely matched the actual values. Local lockdown with social distancing (LD_SD) was found to be ineffective compared to national lockdown. The ranking of effectiveness of supplementary measures for LD_SD were found to be consistent across national hotspots and local areas. Measure effectiveness were ranked from most effective to least effective: 1) full lockdown; 2) LD_SD with international travel −50%; 3) LD_SD with 100% quarantine; 4) LD_SD with closing school −50%; 5) LD_SD with closing pubs −50%. There were negligible differences observed between LD_SD, LD_SD with −50% food & Accommodation and LD_SD with −50% Retail.

**Conclusions:** The second national lockdown should be followed by measures which are more effective than LD_SD alone. Our model suggests the importance of restrictions on international travel and travel quarantines, thus suggesting that follow-up policies should consist of the combination of LD_SD and a reduction in the number of open airports within close proximity of the hotspot regions. Stricter measures should be placed in terms travel quarantine to increase the impact of this measure. It is also recommended that restrictions should be placed on the number of schools and pubs open.

**Strengths and limitations of this study**

- The proposed Deep Recurrent Reinforced Learning (DRRL)-based model takes into account of both relationships of variables across local authorities and across time, using ideas from reinforcement learning to improve predictions.
- Whilst, predicting the geographical trend in COVID-19 cases based on the simulation of different measures in the UK at both the national and local levels in the UK has proved challenging, this study has provided a methodology by which useful predictions and simulations can be obtained.
- The Office for National Statistics only released data on UK international travel up to March 2019 at the time of this study, and therefore this study used the amount of UK tourists in Spain as a reference variable for understanding the effect of international travel on COVID-19 spread.

## INTRODUCTION

COVID-19 is a highly infectious disease that resulted in a global pandemic in just under a month. This pandemic has caused global disruptions to individuals, businesses and governments worldwide. The number of cases have continued to rise exponentially, from 80,239 in February 2020 rising to 69 million as of December 2020.[1] Recent cases of a new variants of COVID-19 have also been found [2].This is despite global efforts to control this virus. Thus, novel strategies are necessary to monitor and control the spread of this virus.

Whilst there have been studies of the geographical distribution of actual COVID-19 cases[3], to the best of our knowledge, there have not been any studies predicting the geographical trend in COVID-19 cases based on the simulation of different interventions at both the national and local authority(LA) levels in the UK. It is vital to have a detailed understanding of the factors that presently affect the spread of COVID-19 at both a national and local level as well as the potential impact of future policy measures. This knowledge would allow the government, LAs and individual citizens to make informed decisions about regional policies and personal exposure risks.

To this end, we have developed a deep learning model that provides the predictions of the incidence and mortality related to COVID-19 in the upcoming 5 weeks and simulates the effect of policy changes targeting the COVID-19 spread i.e. number of facilities available for accommodation and food, pubs, retail shops, education, transport and storage, art, entertainment and recreational services, within each local authority region. The model also accounts for international migration inflow, internal migration inflow and outflow within the UK thus simulating policy changes that affect travel. This model may also inform planning for similar scenarios in the future.

## METHODS

### Patient and public involvement

This research was done without patient and public involvement.

### Model Development

We developed a Deep Recurrent Reinforced Learning (DRRL)-based model (supplementary material Part I) that combines the synergistic properties of Gated Recurrent Units (GRU),[4] and reinforcement deep learning.[5] We have chosen GRU as an element of our model because of its ability to model non-linear and temporal relationships between and within high dimensions of variables. The reinforcement learning element of DRRL enables it to adapt to newly inputted data and make more accurate forecasts.

All available LA data was split 80:20 into training and validation data subsets. Data was pre-processed using scaling - subtracting their corresponding mean and dividing by the standard deviation values. Following the completion of predictions, the prediction outputs are then scaled back to their original scale.

The DRRL neural network model utilised an input layer, numerous hidden layers, and an output layer. A complex series of non-linear matrix computations are applied to the input data to relate the target output (i.e. cases and mortality) to the other data columns (e.g. amount of international migration inflow or internal migration inflow and outflow within UK, number of retail shops etc.). The model is first Trained using the existing data subsets. Each column of the data is assigned a specific weight at each of the nodes in the hidden layers and these weights are progressively updated to minimize the the mean absolute error (MAE) between the predicted and actual values, using the rmsprop optimisation algorithm.[6] During the Prediction process, an input data matrix of the same dimension as the training data is then passed into the input layer. The neural network’s hidden layers then use the weights learned during training process to predict the most likely incidence and mortality based on the values of the other variables from each corresponding week.

The final model consists of two components, model-M (master model) and model-R (reinforced model) that serve different purposes. Model-M accounts for the relationships of variables across different LAs, whilst model-R provides improved forecasting performance for each individual LA that are selected for analysis (see Supplementary Material, Part I). This model is particularly apt at generalisation and is capable of forecasting a wide range of LA simultaneously. The model uses model-R to increase forecast performance for the individual LA that are selected for analysis. Model-M is updated with several additional epochs of training data from the selected LA to reinforce and optimise the predictions.

### Data Linkage

The data used to train the deep learning model is based on various datasets that have the potential to influence the trend in the number of COVID-19 cases and mortality at the national and local level (refer to Supplementary Materials, Part IV. for more details), including domains of deprivation, number of bars and pubs, business size, population estimate (male, female, by age, overall), etc. We use the R language and the R SDK for COVID-19, i.e. a set of software commands to retrieve data remotely, as published by Public Health England, to automatically extract the latest daily cases and mortality figures for all LA within the UK. Using this approach, we are able to automate and dynamically predict the cases and mortality as new data is generated by GOV.UK. We use R to convert these data from daily figures into weekly counts and link this data to the data described in Supplementary Materials, Part IV.

Specifically, we have created a manually curated datasheet containing three indices that together we name the COVID-19 General Policy (CvdGPlc) indices: LockdownScore, QuarantineMeasures and SchoolOpening. These index scores are linked to the main dataset based on the weeks each of the corresponding policies were implemented and the relative effects at each time period.

Furthermore, we obtained the number of tourists arriving in Spain from Jan 2020 to July 2020 and adjusted the number by the proportion of UK tourists in Spain from the year 2019. As data on international travel is not readily available for the period affected by COVID-19, the rationale is to use the amount of UK travel to Spain as an indicator for the impact of international travel on the spread of COVID-19, since Spain is a frequent UK tourist destination. Our GRU model is not only trained on the above data, but also includes the longitude and latitude of each local authority as part of the model.

In this study, we have explored the effects of various containing measures at specific time periods to investigate their effects on virus spread and mortality rates. Only the variables that we have found most relevant as measures for policy making have been reported. The reader is encouraged to explore the adjustment effects of other variables on the prediction results of the model via our online web app (http://137.222.198.54:8081/).

Whilst the LA boundaries data is not included in the training process, the main dataset is also linked to this data following forecast generation, so the deep learning model will also provide the prediction of the incidence or mortality in the next five weeks in a geographical map view. Furthermore, the model has the capability to toggle the map view by local authority or Public Health England regions. These views will not only be useful for the government to see the future effects of different policy changes, but also for the individual citizens to understand their risk of movement within and between local regions in the upcoming future.

For analysis in the map view, the geographical regions from the top to bottom of England is divided into four equidistant slices, which we shall name slice n2, n1, s1, s2, respectively. These categories will be applied to all other geographical plots hereafter to facilitate discussion. The areas with higher number of cases are shown in darker colours with 6 grades of severity (I – VI) covering the ranges 0-250 (I); 250-500 (II); 500-750 (III); 750-1000 (IV); 1000-1250 (V); 1250-1500 (VI). Any number outside of this range is shown in grey and is classed as grade VII.

### Model Validation

The model is validated for the whole of England, whereby the model is trained using the same approach as described in the Model Development section but excluding data from weeks 41 to 46. The data from this interval serves as Validation data for determining the performance of the model on unseen data. Ten iterations of reinforced training are performed over the dataset. At the time of this work, only data up to week 46 are available. The variable values are set from week 40 onwards to enable predictions to simulate a full/national lockdown (FLD) from week 45 onwards. This is because it is known that a FLD had been applied in the UK from week 45 (00.01 on Thursday 5 November) and prior to that, local lockdown with social distancing (LD_SD) had been implemented.

### Model Simulation

Simulations are performed using the final model that is trained using the approach described in the Model Development section. All data i.e. from week 1 to 46 are included for training this model. The model is used to simulate the effects of numerous different COVID-19 prevention measures on the number of cases at week 51 i.e. 5 weeks ahead of the latest available data. The values of the variables that model the corresponding measures are set from week 40 onwards to enable predictions to simulate the implementation of those measures from week 45 onwards, rather than FLD, which was what the government actually implemented. The measures simulated are: (a) No lockdown vs. local lockdown with social distancing (LD_SD); b) LD_SD vs. full/national lockdown (FLD); c) LD_SD vs. LD_SD with international travel −50%; d) LD_SD vs. LD_SD with closing school −50% e) LD_SD with travel quarantine 5.5 (see Supplementary Material, Part IV., 11) vs. LD_SD with full travel quarantine 10; f) LD_SD with 100% pubs open vs. LD_SD with −50% pubs; (g) LD_SD with 100% food & accommodation services open vs. LD_SD with −50% food & accommodation services open; (h) LD_SD with −50% retail services open vs. LD_SD with 100% retail services open. For details on the implementation of these measures, please refer to Supplementary Materials, Part IV.

These measures are simulated firstly for individual LA by selecting a baseline LA with a relatively low case count and comparing the effect of the measures when applied to a LA with a very high number of cases i.e. a hotspot area. The measures are then ranked by order of effectiveness. This is so that the relative effectiveness of each measure can be understood at the local level. Secondly, the measures are simulated for all the LA in England to visualise the relative effectiveness of each measure at a national level. For the 21 LA with the highest cases when using a LD_SD measure, the predicted cases counts at week 51 are extracted and plotted to analyse the efficacy of each measure across these nationally “hard” to tackle areas. This comparison also enabled the ranking of the relative effectiveness of each measure at these hotspots.

## RESULTS

Model validation of predictions against actual results for week 46 showed a good match between the simulation and actual number of cases across all the LA concerned (fig 1). The model was able to distinguish LA with high cases from areas with low number of cases (fig 2a, b). The model performs especially well for low grade LA (Table 1). The tendency towards better performance in low degree LA, may be because data from week 41 to 46 containing sharp changes in the trend have not been included. Therefore, the simulation of cases and mortality up to week 51 was performed by including data from week 41 to 46 in the model training.

**Table 1.** Validation model: Number of actual and predicted cases and mortalities. The results show that there is a close match between actual and predicted number of cases, especially for LA at grade III or below.

| Local Authority | Number of Actual Cases for week 46 | Cases Forecast for week 46 | Number of Actual Mortalities for week 46 | Mortality Forecast for week 46 |
| --- | --- | --- | --- | --- |
| Intervention | Full Lockdown |  |  |  |
| Wolverhampton | 438 | 482 | 4 | 5 |
| Gedling | 179 | 196 | 6 | 2 |
| Welwyn Hatfield | 119 | 130 | 0 | 2 |
| Wiltshire | 201 | 219 | 1 | 4 |
| Portsmouth | 220 | 239 | 0 | 3 |
| Bromley | 217 | 232 | 2 | 3 |
| Stockton-on-Tees | 467 | 498 | 7 | 7 |
| Stockport | 517 | 550 | 13 | 8 |
| South Kesteven | 153 | 162 | 6 | 1 |
| Hammersmith and Fulham | 166 | 175 | 1 | 2 |
| Kingston upon Thames | 150 | 158 | 4 | 2 |
| Ribble Valley | 93 | 98 | 4 | 2 |
| East Cambridgeshire | 34 | 36 | 1 | 1 |
| Redcar and Cleveland | 380 | 396 | 7 | 4 |
| Sedgemoor | 55 | 57 | 3 | 1 |
| Cheshire East | 496 | 514 | 6 | 7 |
| Wealden | 76 | 79 | 1 | 2 |
| Charnwood | 371 | 382 | 3 | 3 |
| South Somerset | 72 | 74 | 1 | 1 |
| Southend-on-Sea | 137 | 140 | 0 | 3 |
| Chelmsford | 110 | 112 | 2 | 2 |
| Rushcliffe | 124 | 126 | 4 | 2 |
| Merton | 146 | 148 | 0 | 2 |
| Shropshire | 426 | 428 | 6 | 6 |
| Harrogate | 253 | 253 | 1 | 2 |
| Central Bedfordshire | 226 | 225 | 5 | 4 |
| Sutton | 155 | 154 | 5 | 3 |
| Oldham | 735 | 732 | 15 | 8 |
| Hillingdon | 325 | 323 | 3 | 3 |
| Basildon | 168 | 167 | 4 | 3 |
| Plymouth | 196 | 192 | 2 | 3 |
| Test Valley | 59 | 58 | 1 | 1 |
| Walsall | 605 | 590 | 15 | 6 |
| Southampton | 187 | 182 | 0 | 2 |
| Selby | 129 | 124 | 2 | 1 |
| South Holland | 105 | 100 | 2 | 1 |
| Chiltern | 57 | 54 | 0 | 1 |
| Derbyshire Dales | 82 | 78 | 1 | 2 |
| Chichester | 58 | 54 | 0 | 1 |
| Barnet | 378 | 354 | 5 | 4 |
| Tameside | 447 | 417 | 18 | 12 |
| Salford | 577 | 537 | 17 | 7 |
| Havant | 77 | 71 | 1 | 1 |
| Waverley | 97 | 89 | 0 | 1 |
| Nuneaton and Bedworth | 247 | 226 | 4 | 3 |
| New Forest | 92 | 84 | 6 | 1 |
| Ryedale | 67 | 61 | 1 | 1 |
| Peterborough | 224 | 204 | 2 | 4 |
| North Hertfordshire | 89 | 81 | 1 | 1 |
| Epping Forest | 130 | 118 | 2 | 1 |

Note: only 50 LA are displayed. For validation data on all LA, please contact the authors.

**Fig 1.**
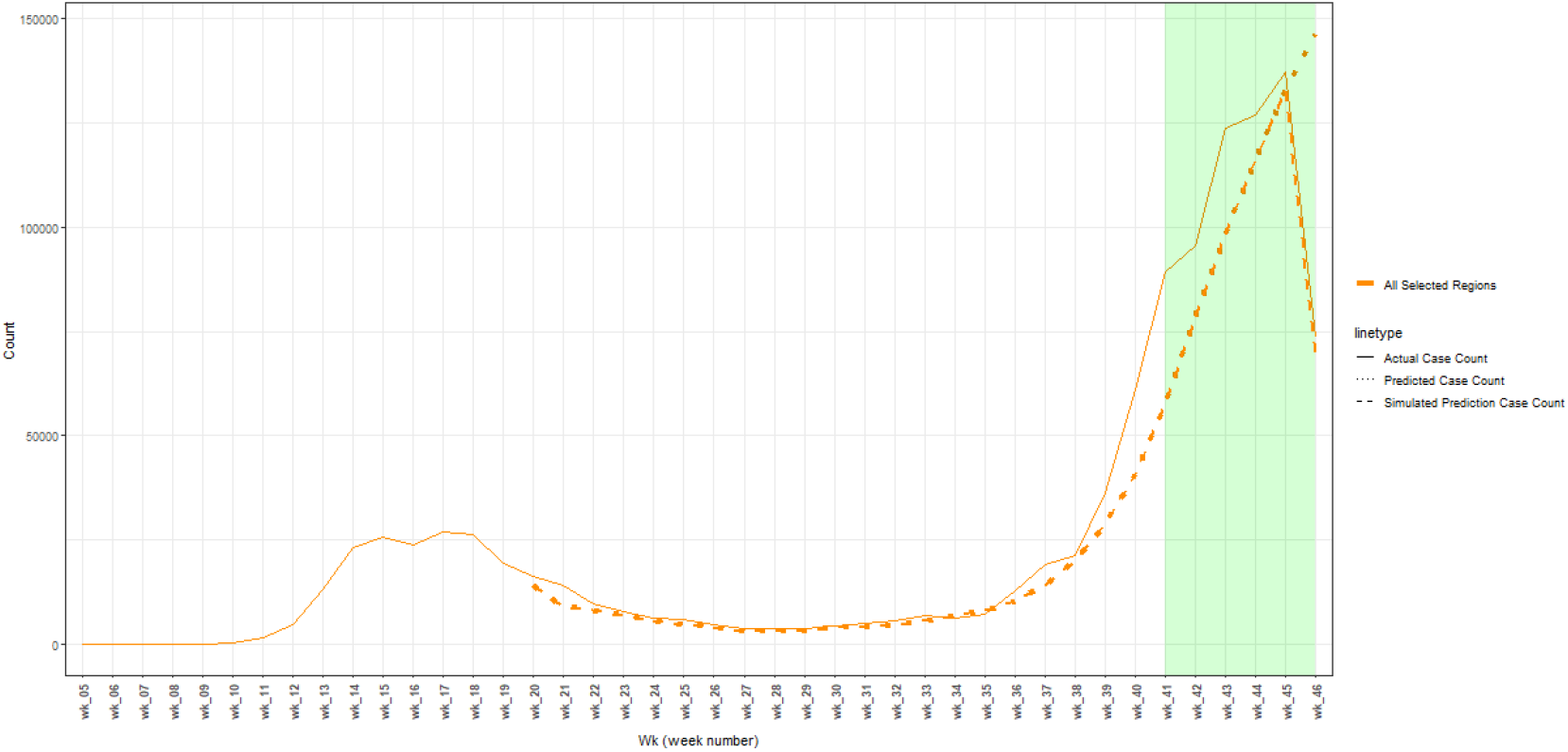
Validation of cases for week 46 with weeks 41 to 46 excluded from data

**Fig 2.**
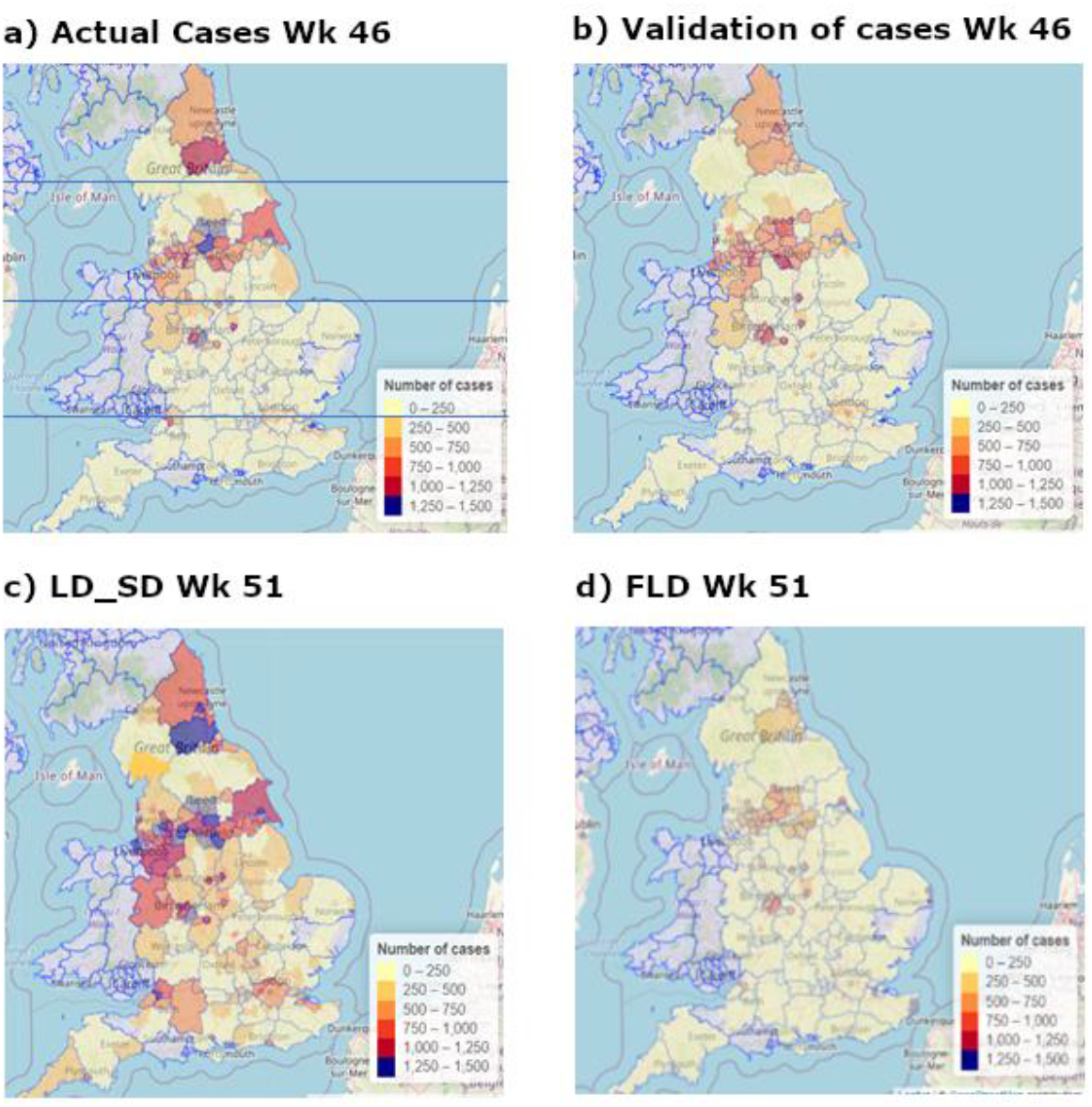
Geographical level of cases for actual and predicted results based on different measures. a) exemplifies the use of geographical slices n2, n1, s1, s2. Additional results are available in Supplementary Materials, Part III.

The effects of different measures were first observed at a local level. Southampton was selected as baseline for observing the effects of measure changes. As Southampton is a grade I LA with a low case number of 187 in week 46, the effects of measure changes were readily perceived with effectiveness ranked from most effective to least effective (Supplementary Materials, Part II fig. S5): b) full lockdown; c) LD_SD & international travel −50%; e) LD_SD & 100% quarantine; d) LD_SD & closing school −50%; f) LD_SD & closing pubs −50%. There were negligible differences observed between LD_SD, g) LD_SD & −50% food & Accommodation and h) LD_SD & −50% Retail.

As Leeds was in the highest grade (VII) for both week 46 (actual) and week 51 (predicted), it was selected for observing the effects of different measures on ‘hard’ to tackle areas. As the number of cases for Leeds were approximately 5 times higher than Southampton, the effect of measures relative to the number of cases in any week were much smaller in the former compared to the latter. For Leeds, no difference was observed for predicted cases at week 51 between no lockdown and LD_SD. Full lockdown (Supplementary Materials, Part II. fig. S6b) was most effective followed by LD_SD with a reduction in international travel by 50%, although the effects were much less in proportion to the number of cases than Southampton. There was negligible impact on the number of cases at week 51 for the remaining measures (fig. S6e-h).

Figure 2c shows the predicted cases in week 51 using LD_SD. At a national level, it can be seen that there would be a rapid rise in the number of cases, especially in the horizontal “belt” along the n1 region. In addition, there is at least one LA in each of the other slices n2, s1 and s2 that are expected to rise to grade VI or above. The majority of LA locations elsewhere, which were mostly at grade I in week 46, are expected to rise to grade II or III. The top 21 hotspots at week 51 using LD_SD were selected for subsequent analysis (Table 2).

**Table 2.** Final model: number of actual and predicted cases and mortalities. Results are shown for the top 21 LA with the highest predicted cases observed at wk 51 using LD_SD.

| Local Authority | Number of Actual Cases for week 46 | Number of Actual Mortalities for week 46 | Cases Forecast for week 51 | Mortality Forecast for week 51 | Cases Forecast for week 51 | Mortality Forecast for week 51 |
| --- | --- | --- | --- | --- | --- | --- |
| Intervention | *Full Lockdown* |  | *Local lockdown with social distancing* |  | *Full lockdown* |  |
| Leeds | 1801 | 17 | 1881 | 21 | 499 | 17 |
| Sheffield | 948 | 36 | 1784 | 32 | 275 | 14 |
| Birmingham | 1957 | 35 | 1627 | 25 | 537 | 17 |
| Wigan | 759 | 34 | 1554 | 24 | 346 | 10 |
| Manchester | 1067 | 13 | 1550 | 20 | 427 | 13 |
| Bradford | 1534 | 24 | 1549 | 20 | 722 | 17 |
| Stockport | 517 | 13 | 1529 | 17 | 218 | 7 |
| Liverpool | 750 | 29 | 1509 | 22 | 234 | 8 |
| Rotherham | 561 | 20 | 1448 | 22 | 257 | 7 |
| Kingston upon Hull | 1011 | 21 | 1368 | 18 | 584 | 12 |
| Oldham | 735 | 15 | 1336 | 17 | 509 | 11 |
| Wirral | 311 | 13 | 1324 | 19 | 65 | 4 |
| Bolton | 635 | 18 | 1299 | 17 | 447 | 11 |
| Bristol | 763 | 8 | 1296 | 14 | 444 | 13 |
| Tameside | 447 | 18 | 1288 | 20 | 255 | 7 |
| County Durham | 1161 | 23 | 1252 | 26 | 377 | 7 |
| Derby | 537 | 11 | 1234 | 15 | 335 | 8 |
| Walsall | 605 | 15 | 1233 | 21 | 288 | 7 |
| Kirklees | 1292 | 25 | 1207 | 29 | 557 | 14 |
| Leicester | 1006 | 7 | 993 | 15 | 581 | 11 |
| Sandwell | 762 | 21 | 969 | 25 | 398 | 10 |

LD_SD was shown (fig 3) to be effective in suppressing the increase in cases for Birmingham (−17%), Bradford (+0.98%), Kirklees (−6.6%) and Leicester (−1.3%). LD_SD was shown to be ineffective for suppressing the increase in cases for the remaining 17 LA, with the highest predicted rises for Wirral (325%), Stockport (163%), Tameside (188%), Rotherham (158%), Derby (130%).

**Fig 3.**
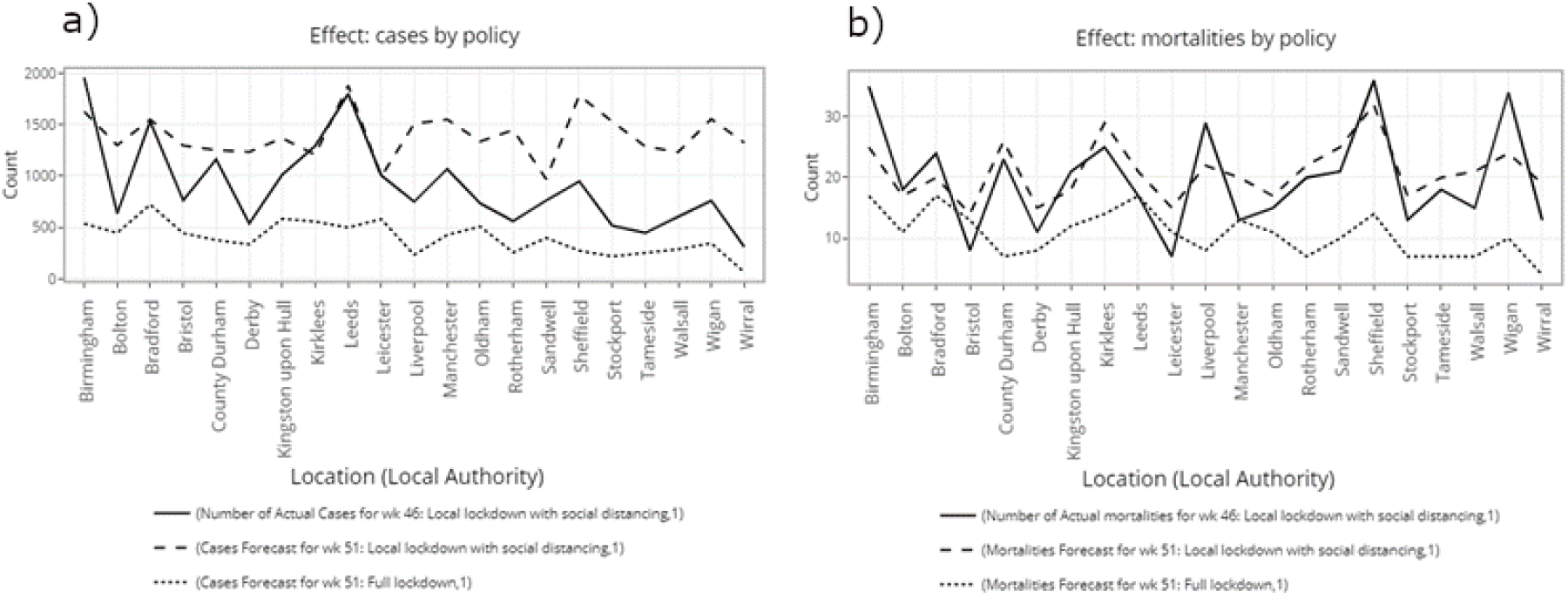
For the top 21 LA with the highest predicted cases observed at wk 51 using LD_SD, plots were generated to compare the effects of full lockdown against LD_SD in terms of cases a) and mortalities b).

LD_SD with −50% international travel was the most effective measure after full lockdown (blue vs. brown, fig 4). 100% quarantine (pink) was the next most effective supplementary measure, with similar effectiveness to international travel −50% except for three LA. Notably, LD_SD with 100% quarantine resulted in higher cases than LD_SD with international travel −50% for Bradford (+9.1%), Leicester (+7.6%). As an exception, Manchester had -41% less cases when using the quarantine measure compared to international travel restrictions.

**Fig 4.**
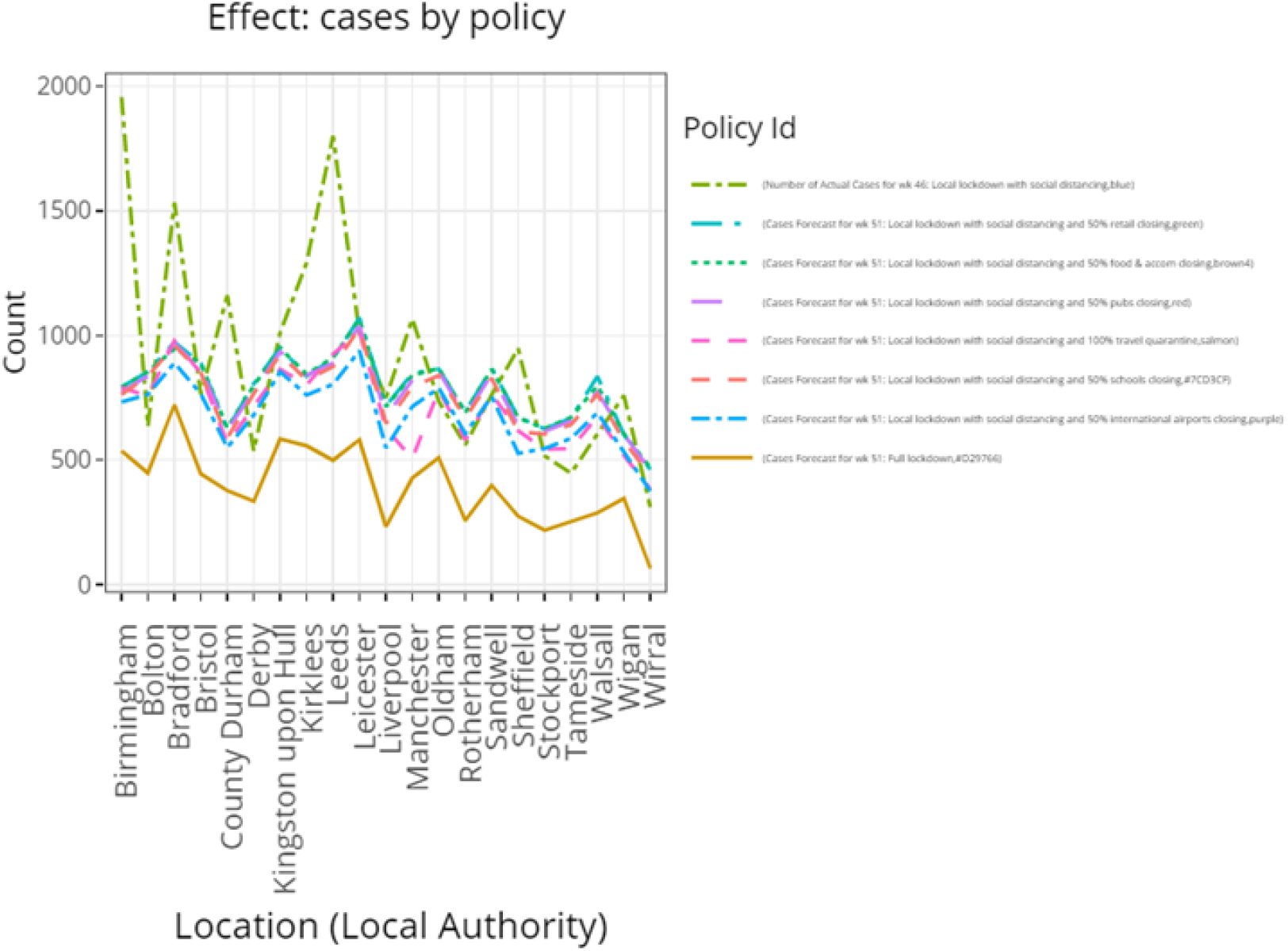
For the top 21 LA with the highest predicted cases observed at wk 51 using LD_SD, a plot is generated to compare the effect on the number of cases using a combination of LD_SD with other “supplementary” measures.

The supplementary effect of school closing −50% was less than international travel restrictions for all 21 LA, with the number of cases being (+9.2%) higher on average using the former measure. Closing pubs −50% had a similar, albeit slightly lower level of effectiveness compared to school closing, with a higher number of cases (+2.2%) on average using the former measure compared to the latter. Again, reducing the number of food & accommodation services −50% had a similar, but slightly lower level of effectiveness compared to pubs closing, with the number of cases (+2.0%) being higher on average using the former measure. In addition, a reduction in the number of retail services −50% resulted in a similar effect to food & accommodation services −50%, with on average a minimal increase in the number of cases (+0.29%) using the former measure. It can be seen that on average, the ranking of measure effectiveness for the national hotspots are the same as the local baseline, i.e. Southampton.

## DISCUSSION

We have developed a deep learning model that investigates the impact of local versus national measures on COVID-19 spread in England as well as the associated mortality rates and allows forecasting based on simulation of different scenarios (http://137.222.198.54:8081/). This model can be regularly updated as the new information on actual numbers becomes available.

The temporal based deep learning model can be used to make inferences into the effectiveness of different measures at both the national and local level. The model suggests that there is variation in the effect of each measure across different regions. Notably, our results suggest that the protective effects of lockdown measures benefit some local authorities more than others (Supplementary Material, Part II. fig S5 and S6) and that local lockdown with social distancing is ineffective compared to national lockdown in suppressing the increase in cases for most of the local authority areas. That is, if the government had kept the same local lockdown with social distancing policies, which they had implemented from week 40 onwards rather than switching to a national lockdown policy at week 45, then we would have seen a rapid rise in cases not only in the n1 belt region, but also in areas such as County Durham (n2), Bristol (s2) and Birmingham (s1), as well as in many other areas across England.

Local lockdown with social distancing may be inefficient in stopping rapid rise of hotpot regions due to geographical properties of hotspot regions. Hotspots along the middle of the n1 geographical slice constitute a tight cluster of large metropolitan cities, and the high number of cases may be partly attributed to the high number of services such as pubs and schools available as well as the amount of travel in these areas. We also expect that LA areas where there are many boundary connections to other hotspots are likely to develop a higher number of cases when LD_SD measures are implemented. This is because, if one of an LA’s neighbours is locked down, citizens from that LA can still travel to its other neighbours given that it has many neighbouring connections. We expect this could allow the continuation of the spread of COVID-19 within these tight cluster regions.

Since the government is only able to impose a national lockdown for a limited period, follow-up measures should improve upon LD_SD as this is likely not to be sufficient. The introduction of additional measures on top of local lockdown with social distancing can help to suppress the increase of or even decrease the number of cases in national hotspots as well as local areas where cases are not very high. Our model shows that the ranking of the average effectiveness of each supplementary measure is consistent across the national hotspots and local baseline, and this ranking can be used to prioritise those interventions according to an order of effectiveness. Nonetheless, it was also observed that certain measures are more effective for some LA compared to others. In these cases, it is necessary to adjust the priorities of the measures implemented accordingly.

The model has highlighted the importance of reducing the amount of international travel, the number of open schools and pubs as well as the implementation of travel quarantine procedures in controlling the spread of COVID-19 over other measures, such as reducing the number of food & accommodation and retail services, which seemed less relevant on the virus spread (fig 4 and fig S5). One explanation for the importance of international travel on the spread of the disease is that whilst the UK government has a certain level of control over the restriction of activities in the UK, it has little control over the activities of travelling individuals once they arrive at their destinations as well as the level of health preventative measures at those destinations. Furthermore, the travelling individuals are more likely to encounter hotels, resorts, trains, planes and other places of gathering whilst abroad. These factors can contribute to the increased spread of COVID-19. Whilst travel quarantine measures provide the government to some degree the selection of which countries to enforce a 14-day self-isolation on the traveller’s return, there are possible explanations why this would not be as effective as directly reducing the amount of international travel. Firstly, the restrictions do not prevent travellers (e.g. pre-university gap year students, those not working) not wary of self-isolation to travel to high risk countries. Secondly, although a penalty is imposed if self-isolation is violated, the act of self-isolation is largely dependent on the level of cooperation from the individual. Finally, even with COVID-19 testing in place, the journey from the airport back to the home of the traveller allows an increased opportunity of spreading the virus, particularly if public transport such as taxis or buses are taken.

Whilst closing schools were not as effective as international travel and quarantine restrictions, it was found to be more effective than closing pubs. One potential explanation for this is that schools are more crowded places and are subject to more frequent number of close contact scenarios in comparison to the pub. The view that schools contribute to the spread of COVID-19 have been supported by the literature.[7,8] Whilst the virus may pose low risk of mortality to the children themselves, these frequently asymptomatic carriers can also lead to the spread of the virus to their households, teachers and communities.

The reason why minimal effects were found for food & accommodation and retail restrictions may be due to the possibility that in these sectors people generally associate with others that they are closely associated with. For example, families are more likely to sit with each other in restaurants or walk together with each other when shopping rather than people they are less familiar with. This is not the case in pubs as anyone from the communal area can be present.

It is unexpected that in the s2 slice that Bristol has higher predicted cases than the LA in the London area as one would have thought the latter comprising a total population of 9 million (2019) and a high traffic volume owing to its large underground network system would result in much higher case numbers. We expect that this may be because the LA in the London region generally have less health and disability deprivation (Deciles: Wandworth: 7; Barnet: 8.9; Brent: 7.3; Waltham Forest: 6.1) compared to Bristol (Decile: 4.4). This is supported by the finds which suggest that existing comorbidities are associated with an increased likelihood of COVID-19 hospital admission.[9]

In light of evidence given by the comparison between the LA within the London region and Bristol, we expect the effect of LA boundary connections to be adjusted by the degree of health and disability deprivation. Indeed, we found that regions with high number of cases along the horizontal “belt” in the n1 region had a high degree of health and disability deprivation (Decile: Manchester: 1.9; Leeds: 4.1; Bradford: 3.3; Liverpool: 1.8; Sheffield: 3.9; Wigan: 3.4). This also applies to County Durham (n2), which has a high degree of health and disability deprivation (decile: 2.9) and was seen to have a significant increase in cases at week 51 using a LD_SD measure.

## CONCLUSION

The present analysis found that the national lockdown was more effective than targeted local lockdown measures and its implementation from week 45 to 49 by the government was justified, albeit not sufficient to fully control the spread of COVID-19. In addition, given the limited governmental resources and timespan of the national lockdown, a rapid rise would be inevitable in health deprived and geographically vulnerable areas with a high degree of boundary connection, if only local lockdown with social distancing is used as a followed-up measure.

Our model suggests the importance of restrictions on international travel and travel quarantines, thus suggesting that follow-up policies should be comprised of the combination of local lockdown with social distancing and a reduction in the number of open airports within close proximity of the hotspot regions. Stricter measures should be placed in terms travel quarantine to increase the impact of this measure. In addition, it is recommended that where possible, education should be provided remotely, and pubs should be closed.

## Data Availability

All the data used in the study are available from public resources. The data source are found in the Supplementary Materials document and any study results can also be made available on request from the corresponding author.

## FOOTNOTES

### Contributors

TD and UB conceived and designed this study. TD and UB acquired the data. TD, UB, SS, and AD analysed and interpreted the data. MC and GDA provided administrative and operational support. The initial manuscript was drafted by TD; all authors critically revised the manuscript and approved its final version. UB acts as guarantor. The corresponding author attests that all listed authors meet authorship criteria and that no others meeting the criteria have been omitted.

### Copyright/license

*I, the Submitting Author has the right to grant and does grant on behalf of all authors of the Work (as defined in the below author licence), an exclusive licence and/or a non-exclusive licence for contributions from authors who are: i) UK Crown employees; ii) where BMJ has agreed a CC-BY licence shall apply, and/or iii) in accordance with the terms applicable for US Federal Government officers or employees acting as part of their official duties; on a worldwide, perpetual, irrevocable, royalty-free basis to BMJ Publishing Group Ltd (“BMJ”) its licensees and where the relevant Journal is co-owned by BMJ to the co-owners of the Journal, to publish the Work in BMJ Open and any other BMJ products and to exploit all rights, as set out in our [licence](https://authors.bmj.com/wp-content/uploads/2020/09/BMJ-Journals-Combined-Author-Licence-November-2018.pdf)*.

*The Submitting Author accepts and understands that any supply made under these terms is made by BMJ to the Submitting Author unless you are acting as an employee on behalf of your employer or a postgraduate student of an affiliated institution which is paying any applicable article publishing charge (“APC”) for Open Access articles. Where the Submitting Author wishes to make the Work available on an Open Access basis (and intends to pay the relevant APC), the terms of reuse of such Open Access shall be governed by a Creative Commons licence – details of these licences and which [Creative Commons](https://creativecommons.org/) licence will apply to this Work are set out in our licence referred to above*.

## Funding

This study was supported by the NIHR Biomedical Research Centre at University Hospitals Bristol and Weston NHS Foundation Trust and the University of Bristol. The funders had no role in the study design, data collection and analysis, decision to publish, or preparation of the manuscript. All authors had full access to all of the data (including statistical reports and tables) in the study and can take responsibility for the integrity of the data and the accuracy of the data analysis.

## Competing interests

All authors have completed the [Unified Competing Interest form](http://www.icmje.org/coi_disclosure.pdf) (available on request from the corresponding author) and declare: no support from any organisation for the submitted work; no financial relationships with any organisations that might have an interest in the submitted work in the previous three years; no other relationships or activities that could appear to have influenced the submitted work.

## Ethical approval

Not needed

## Transparency

The manuscript’s guarantor affirms that the manuscript is an honest, accurate, and transparent account of the study being reported; that no important aspects of the study have been omitted; and that any discrepancies from the study as planned (and, if relevant, registered) have been explained.

## Data sharing

All the data used in the study are available from public resources. The dataset is found in the Supplementary Materials document and can also be made available on request from the corresponding author.

## Notes

### Competing Interest Statement

The authors have declared no competing interest.

### Author Declarations

Approval not required

