## Supplementary material for "A Deep Recurrent Reinforced Learning model to compare the efficacy of targeted local vs. national measures on the spread of COVID-19 in the UK": Figure S1, Figure S1, Figure S1, Figure S4.1,

Part I.

### GRU Gated Recurrent Units Model

We have chosen a Recurrent Neuro Network (RNN) category of model because of the ability of this type of model to model not only non-linear relationships between high dimension of variables, but also because of its ability to model temporal relationships within any of the variables considered. As figure S1. shows, the RNN model consists of cells is analogous to each unit of the traditional neuro network. Whilst traditional neuro network models accumulate weights $W_i$ (input to hidden layer) and $W_h$ (hidden to hidden layer), they do not calculate any recurrent weights $W_r$ that represent temporal relationships of each variable in the dataset. This recurrent type of weight is present in RNN models and hence is desirable for modelling a time dependent COVID-19 dataset.

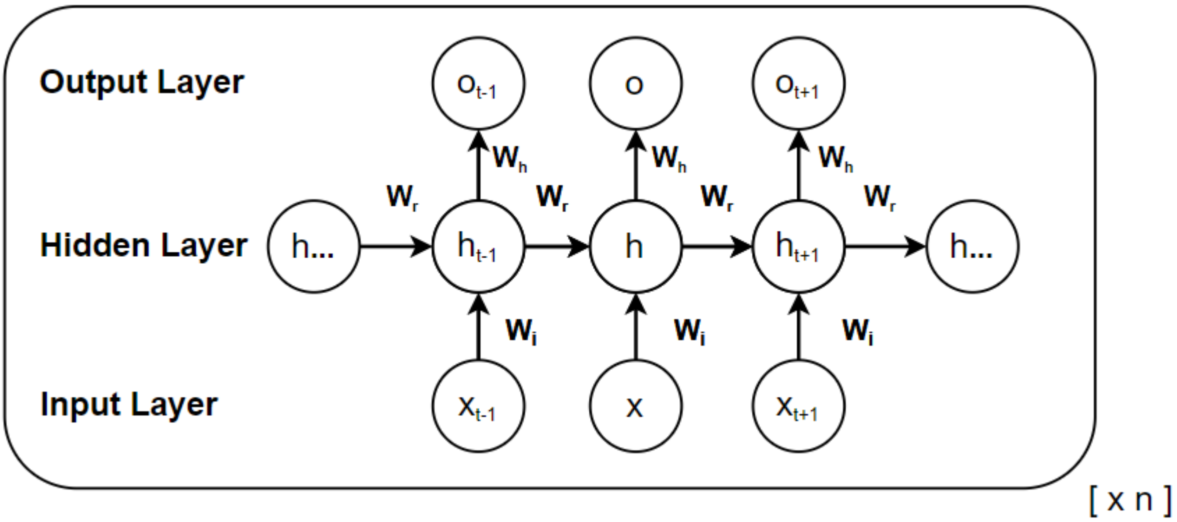


Figure S1.  An RNN cell

GRU is a specific type of Recurrent Neuro Network (RNN) that incorporates long term memory and effectively deals with the vanishing gradient and gradient explosion problems that have affected various other categories of RNN. The GRU model was also selected because of its ease to optimise and its low computational cost in comparison to the LSTM model.

Unlike the LSTM model, which uses four gates, the GRU model uses only a single decision gate that controls both the update and reset of weights. The update gate is similar to an OR gate in electronics that either retains the state at the previous step $h_{t-1}$ or updates the previous state based on variable values $X_t$ provided at the current step.  The reset gate is much like a resistor in that it controls that size of the effect from previous step that contributes to the current step.

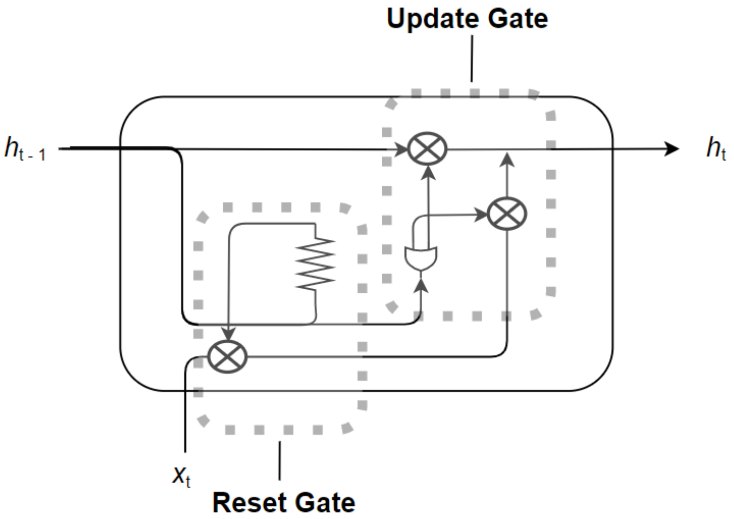

Figure S2.  A GRU cell

The above diagram above provides an intuitive understanding into the processes that occur within a cell or node of a GRU model. The specific mathematical equation of the GRU model is given below:

$$h_t^i = z_t^i . h_{t-1}^i + \left(1 - z_t^i\right) tanh\left(b^i + \sum_j W_I^{i,j} x_t^j + \sum_j W_R^{i,j} r_{t-1}^j h_{t-1}^j\right)$$
$$z_t^i = \sigma\left(b_z^i + \sum_j W_z^{i,j} x_t^j + \sum_j W_z^{i,j} h_{t-1}^j\right)$$
$$r_t^j = \sigma\left(b_r^i + \sum_j W_r^{i,j} x_t^j + \sum_j W_r^{i,j} h_{t-1}^j\right)$$

where z represents for the update gate and r represents the reset gate, $W_I$ are the weights from the input to hidden layer, $W_R$ are the weights from the recurrent weights, b is the  bias, $\sigma$ and tanh are the sigmoid and tanh activation functions respectively, $X_t$ are the inputs at time t and $h_{t-1}$ was the input from previous time step.

**Deep Reinforcement Learning**

Deep reinforcement learning is the application of reinforcement learning to neuro networks. The application of this approach has been exemplified in the field of computer gaming.[1] Essentially, the approach involves an agent that represents the computer system, which performs an action in the environment that it interacts with.

The environment is changed following the action and a reward is provided to the agent such that the action it selects from a list of actions or policy $\pi$ is optimised in any subsequent interactions with the environment.

Here, we present a technique whereby the NCGFS model represents the agent. It is initially presented with a set of observations $O_t$ from the national environment, *s*. Each individual observation has the property o ∈ *A*, where *A* is the set of all the LA. Rather than using the traditional reward variable *r*, we use $L_t$ to represent the loss at the initial training phase *t* of the deep learning model, i.e. at the generation point of model-M. Subsequently, when the agent is presented with the input observations $O_{t+1}$ that belongs to a single local authority, $s_{t+1}$ from the set A, it uses the action or in this case forecast from the list of potential forecasts that previously minimised the $L_t$ for $O_t$ to make the forecast $f_t$ for $O_{t+1}$. This induces a loss $L_{t+1}$ that is used to update the NCGFS model's policy decisions for the actual prediction of $O_{t+1}$ as $f_{t+1}$. The resulting model is termed model-R.

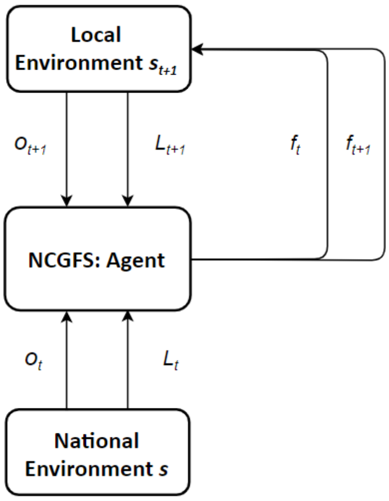


Figure S3. illustration of the Deep Reinforcement Learning aspect of NCGFS at the local authority level.

The following equation shows how NCGFS uses deep reinforcement learning to minimise the Loss $L$ for a given environment s to produce an optimal forecast from the probabilistic distribution of forecasts $\pi = P(f|s)$.

$$Q^*(s,\ f) = \min_{\pi} \frac{1}{n} \left( \sum_{i}^{n} \gamma^i L_{t+i} \mid s_t = s,\ f_t = f,\ \pi \right)$$

Neural Network architecture and configuration

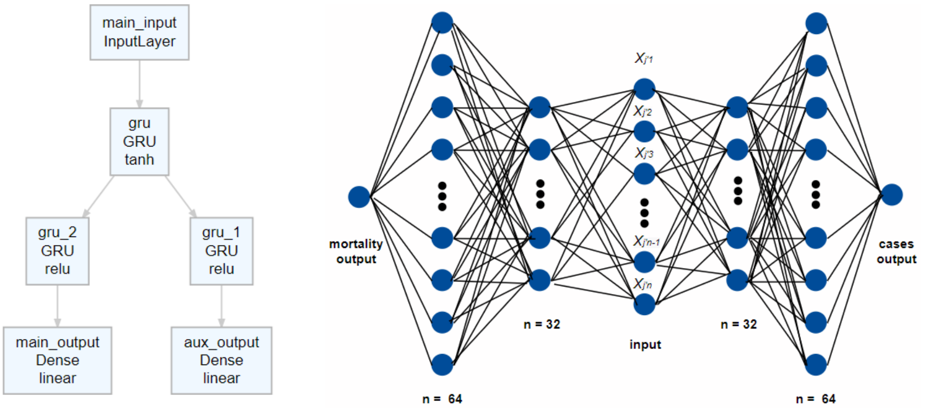


Figure S4.1 NCGFS model architecture. Note, the two layers on either side of the input layer have been depicted in this manner to facilitate representation but represent the same layer.

The model is comprised of symmetrical neuro network that consists of six layers. The input layer accepts a data matrix in the dimension of $[d \quad \tau \quad n]$, where $d$ = 136 is the number of variables, $\tau$ = 2 is the number of time steps in the recurrent direction along the hidden layer, $n$ is the number of samples taken from the observation. The layer immediately right of the input layer is named gru and consists of 32 GRU cells or units. The next layer to the right is name gru_2 and consists of 64 GRU cells. The output from this layer uses the ReLU activation function as this has been demonstrated to be effective for the convergence of neuro networks. This is followed by a dense output layer named main_output, consisting of one unit on the right. This layer provides the prediction output for the number of COVID-19 cases five weeks ahead of the corresponding weeks in the input data.

The layer immediately left of the input layer is in fact the same layer as that to the right of input layer i.e. named gru. The next layer to the left is a named gru_1 and consists of 64 GRU cells. This is followed by a dense output layer named aux_output for prediction the number of mortalities five weeks ahead of the corresponding weeks in the input data.

We have empirically found that this combination of depth and width is efficient for the minimising the loss of the learning problem at hand.

Model-M was trained using four iterations of 500 epochs (with 6 steps per training epoch and 1 step per validation epoch) using early stop to stop training once validation loss has stopped decreasing with a difference of 0.001 and patience of 200. With the four iterations of training, NCGFS model-M had achieved a performance validation loss of 0.17456 consisting of the sum of validation loss for both cases and mortality.

Part II.

1. File 2: domains of deprivation

url location:
https://www.gov.uk/government/statistics/english-indices-of-deprivation-2019
2019 indices containing individual metrics including health, education
Index of Multiple Deprivation (IMD) Decile (where 1 is most deprived 10% of LSOAs)
IMD Rank (1 is most deprived)

2. 2001 to 2018 edition of this dataset

Public houses and bars by local authority

url location:

https://www.ons.gov.uk/businessindustryandtrade/business/activitysizeandlocation/datasets/publichousesandbarsbylocalauthority

Using Pubs size LA 2018 by local authority. Gives the number of pubs in UK by local authority

3. 2020 edition of this dataset

UK business: activity, size and location

url location:

https://www.ons.gov.uk/businessindustryandtrade/business/activitysizeandlocation/datasets/ukbusinessactivitysizeandlocation

ukbusinessworkbook2020.csv by Retail, Transport_Storage_inc_postal, Accommodation_food services, Education, Health, Arts_entertainment recreation_other_services

4. Local Authority Districts 2019 boundaries

Local Authority Districts in the United Kingdom, as at 31 December 2019. The boundaries available are:

- (BUC) Ultra Generalised (500m) - clipped to the coastline (Mean High Water mark).

https://geoportal.statistics.gov.uk/datasets/local-authority-districts-december-2019-boundaries-uk-buc

We use this as COVID-19 cases data is based on 2019 LA boundaries

5. Local Authority District to Public Health England Centre to Public Health England Region (December 2019) Lookup in England

url:

https://geoportal.statistics.gov.uk/datasets/local-authority-district-to-public-health-england-centre-to-public-health-england-region-december-2019-lookup-in-england

6. Mid-2019: April 2019 local authority district codes edition of this dataset

Population estimate data

url:
https://www.ons.gov.uk/peoplepopulationandcommunity/populationandmigration/populationestimates/datasets/populationestimatesforukenglandandwalesscotlandandnorthernireland

Using the following sheets: MYE2 – Males, MYE2 – Females, MYE3 (migrationFlow), MYE2 – Persons (contains age population).

7. SchoolOpening

url:

https://www.gov.uk/government/publications/actions-for-schools-during-the-coronavirus-outbreak/guidance-for-full-opening-schools

https://www.bbc.co.uk/news/uk-51952314

Schools have remained open to some pupils since 23 March, welcoming more pupils back from 1 June.

During model generation, the SchoolOpening index was linked to the main dataset based on the weeks the school restriction policies were implemented and the relative effects at each time period. Index definition: 3 = No restrictions; 1.5 = School closing but remaining open to some pupils; 3 = Also used to represent school reopening.

Unlike the LockdownScore indices, the SchoolOpening index already takes into account of the effect that school closing typically occurs when the number of cases or mortality is highest, by inversing scoring. Hence, in this case when varying the SchoolOpening parameter during model configuration mode, this metric will not have an inverse effect.

Note, the effects pertain only to the local authorities selected.

8. Number of monthly arrivals in tourist accommodation in Spain from August 2018 to July 2020*

url:

https://www.statista.com/statistics/1130775/number-of-monthly-arrivals-short-stay-accommodation-in-spain/

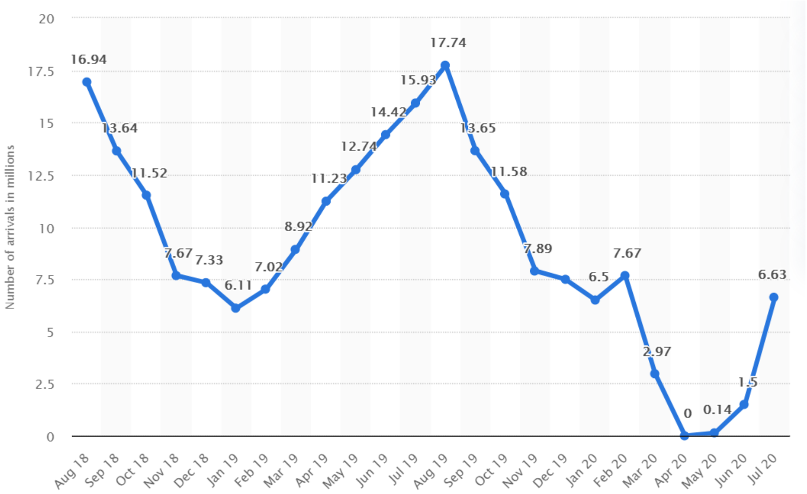


Figure S4.2. Monthly worldwide tourist arrivals in accommodation in Spain

We adjust tourist arrival by the proportion of UK citizens travelling to Spain in 2019

(section 9.)

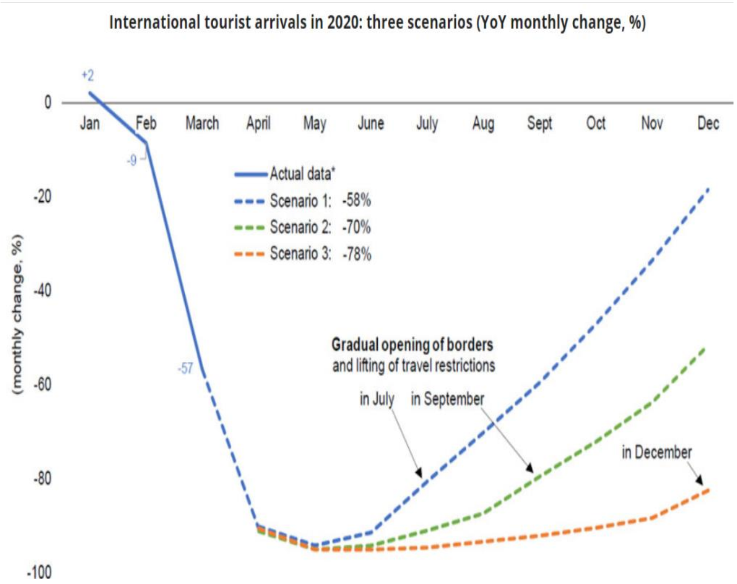


Figure S4.3 A reference model for understanding how quarantine measures are modelled.

url: https://www.adpr.co.uk/blog/covid-19/travel-and-tourism-brands-can-recover-from-coronavirus/

As UK removed 75 countries from quarantine list in July, we impute missing values from August to October based on the recovery model for July opening of borders above. However, the actual recovery based on the data is much faster than that shown in figure above.

9. Number of international tourists arriving in Spain in 2019, by country of residence

url:
https://www.statista.com/statistics/447683/foreign-tourists-visiting-spain-by-country-of-residence/

Obtains proportion of UK citizens arriving in Spain in 2019 as estimate for proportion in 2020

statistic_id447683_international-tourist-arrivals-in-spain-2019-by-country-of-residence.xlsx

UK proportion: 0.215984348

This is the estimated number of tourists arriving in Spain from UK in millions. The values are calculated based on number of tourists from various countries arriving in Spain from Jan 2020 to July 2020 and then adjusting by multiplying this number by the proportion of UK tourists in Spain from the preceding year i.e. 2019.

The weeks not covered by the data available i.e. August to October are imputed based on the tourism industry recovery model for July opening of borders https://www.adpr.co.uk/blog/covid-19/travel-and-tourism-brands-can-recover-from-coronavirus/.

This metric should be used as a guidance for how the situation will vary when international travel is restricted.

10. Public Health England Centres (December 2016) Ultra Generalised Clipped Boundaries in England

url:

https://geoportal.statistics.gov.uk/datasets/public-health-england-centres-december-2016-ultra-generalised-clipped-boundaries-in-england

2016 is the latest available

11. QuarantineMeasures

url:

https://webcache.googleusercontent.com/search?q=cache:J8RTskJUc0QJ:https://www.theweek.co.uk/107044/UK-coronavirus-timeline+&cd=4&hl=en&ct=clnk&gl=uk

During model generation, the QuarantineMeasures index were linked to the main dataset based on the weeks the travel quarantine policies were implemented and the relative effects at each time period. Index definition: 1 = No quarantine; 10 = full quarantine of tourist from all countries; 5 = Removal of 59 countries from quarantine list; 5.5 = Adding Spain back to the quarantine list following removal of 59 countries from the list.

As the level of travel quarantine restrictions implemented is dependent on the severity of covid-19 situation in other countries rather than the number of cases and mortality in UK, when varying this parameter during model configuration mode, this metric will have not an inverse effect.

Note, the effects pertain only to the local authorities selected.

12. LockdownScore

During model generation, the LockdownScore index were linked to the main dataset based on the weeks the lockdown policies were implemented and the relative effects at each time period. Index definition: 1 = No lock down; 4 = Local lock down; 5 = Local lock down with social distancing; 10 = Full Lock Down.

However, as the level of lock down implemented is typically highest when the number of cases or mortality is highest, when varying this parameter during model configuration mode, this metric will have an inverse effect. That is, changing the LockdownScore value to 1 will result in a full lock down, whilst changing to 10 will result in no lock down. 4 will result in Local lock down with social distancing and 5 will represent Local lock down.

Note, the effects pertain only to the local authorities selected.

Part III.

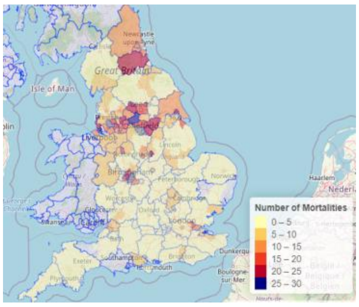


(i) M: Actual Mortalities Wk 46

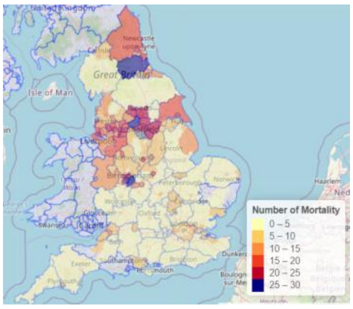


(ii) M: Wk 51 Local LD_SD

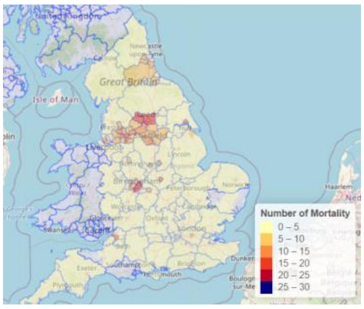


(iii) M: Wk 51 FLD

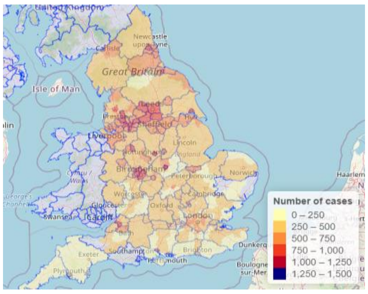


(iv) C: LD_SD -50% school

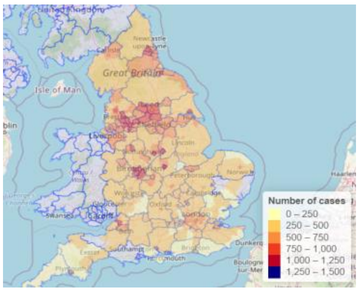


(v) C: LD_SD -50% food & accom

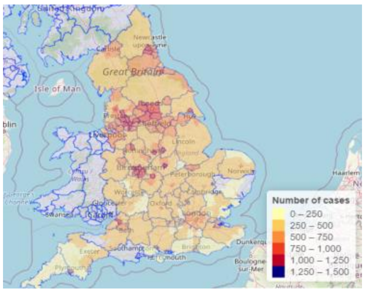


(vi) C: LD_SD -50% retail

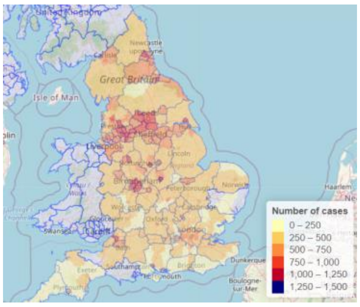


(vii) C: LD_SD -50% pubs

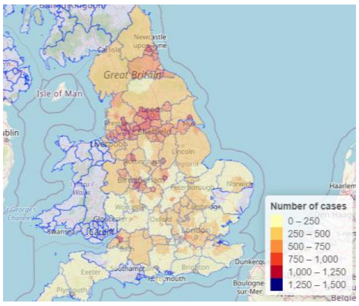


(viii) C: LD_SD intl travel -50%

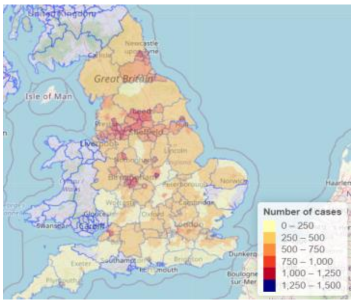


(ix) C: LD_SD 100% quarantine

Figure S5. Geographical level of cases and mortality for actual and predicted results based on different measures. C: cases; M: mortalities.

Part IV.

The deep learning model was used to generate the plots for fig S5. of the effects on the predicted number of cases for Southampton based on different measures.

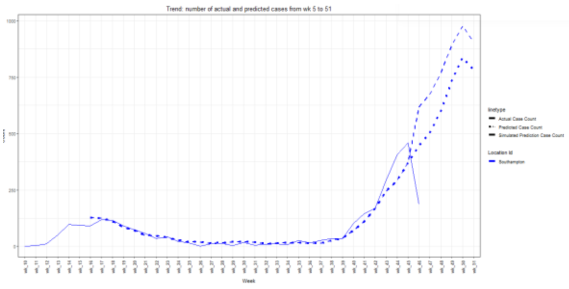

(a) No lockdown vs. LD_SD

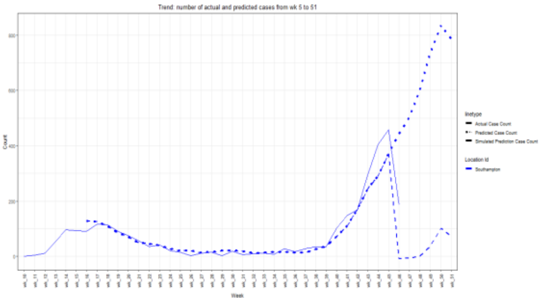

(b) LD_SD vs. full lockdown

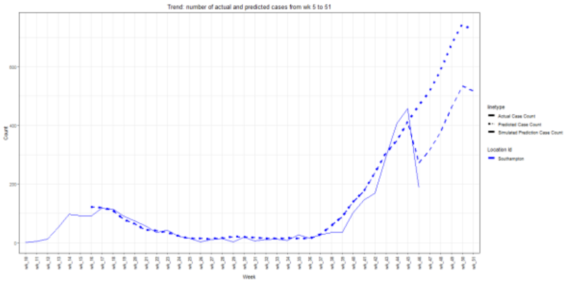

(c) LD_SD vs. international travel -50%

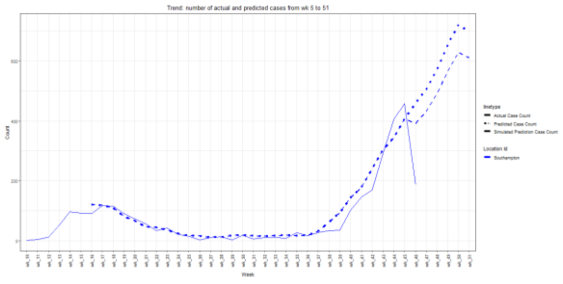

(d) LD_SD vs. closing school -50%

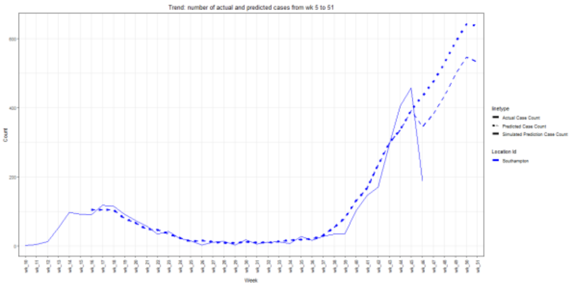


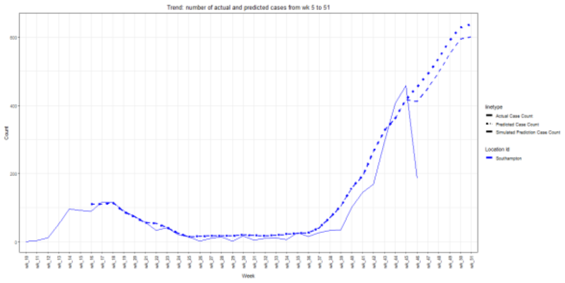


(e) LD_SD (quarantine 5.5) vs. full quarantine (10) (f) LD_SD (100% pubs) vs. -50% pubs

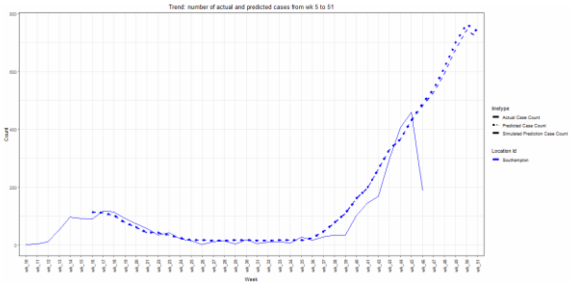


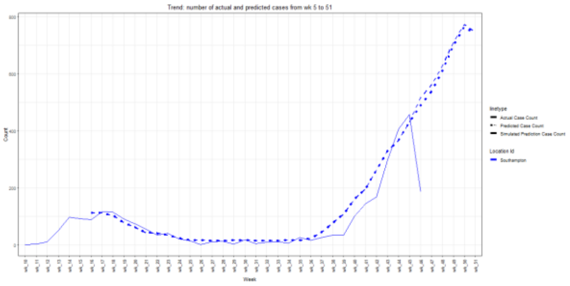


(g) LD_SD (100% food & Accom) vs. -50% (h) LD_SD (100% Retail) vs. -50% Retail

Figure S6. Cases forecast for Southampton by measure. The dotted line represents predictions using the LD_SD measures. The dashed lines represent prediction changes based on changes to measures. (c-h) relate to LD_SD without (left) and with (right) the supplementary measures.

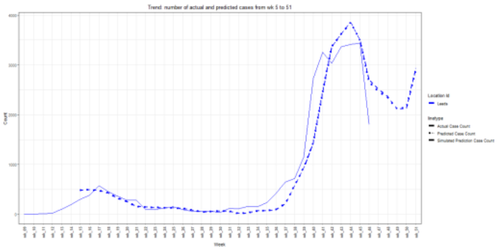


(a) No lockdown vs. LD_SD

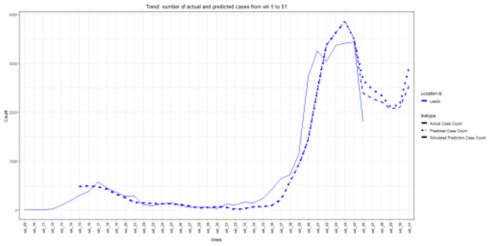


(b) LD_SD vs. full lockdown

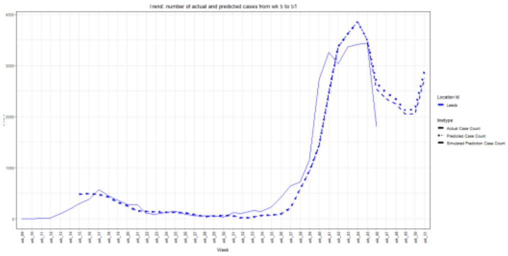


(c) LD_SD vs. international travel -50%

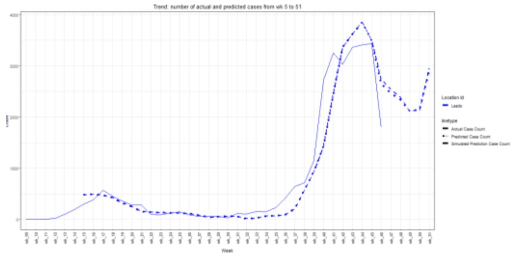


(d) LD_SD vs. closing school -50%

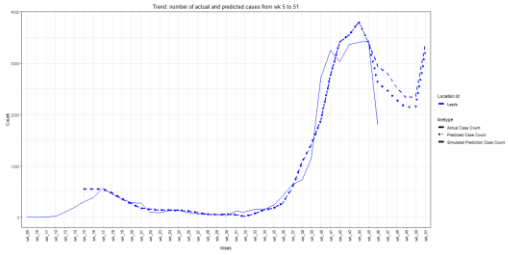


(e) LD_SD (quarantine 5.5) vs. full quarantine (10)

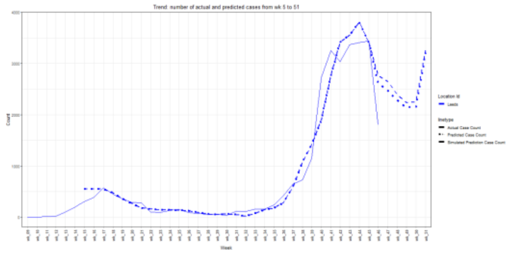


(f) LD_SD (100% pubs) vs. -50% pubs

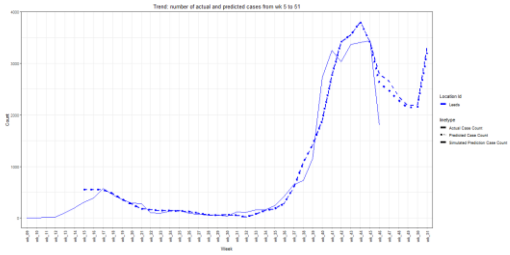


(g) LD_SD (100% food & Accom) vs. -50%

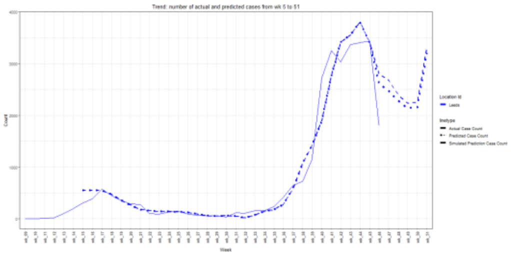


(h) LD_SD (100% Retail) vs. -50% Retail

Figure S7. Cases forecast for Leeds by measures. The dotted line represents predictions using the LD_SD measures. The dashed lines represent prediction changes based on changes to measures. (c-h) relate to LD_SD without (left) and with (right) the supplementary measures.
